# Using deep convolutional neural networks to predict patients age based on ECGs from an independent test cohort

**DOI:** 10.1101/2022.10.03.22280640

**Authors:** Bjørn-Jostein Singstad, Belal Tavashi

**Affiliations:** Simula Research Laboratory; Department of Biomedical Engineering, Ankara University

## Abstract

Electrocardiography is one of the most frequently used methods to evaluate cardiovascular diseases. However, the last decade has shown that deep convolutional neural networks (CNN) can extract information from the electrocardiogram (ECG) that goes beyond traditional diagnostics, such as predicting a persons age. In this study, we trained two different 1-dimensional CNNs on open datasets to predict age from a persons ECG.

The models were trained and validated using 10 seconds long 12-lead ECG records, resampled to 100Hz. 59355 ECGs were used for training and cross-validation, while 21748 ECGs from a separate cohort were used as the test set. We compared the performance achieved on the cross-validation with the performance on the test set. Furthermore, we used cardiologist annotated cardiovascular conditions to categorize the patients in the test set in order to assess whether some cardiac condition leads to greater discrepancies between CNN-predicted age and chronological age.

The best CNN model, using an Inception Time architecture, showed a significant drop in performance, in terms of mean absolute error (MAE), from cross-validation on the training set (7.90 ± 0.04 years) to the performance on the test set (8.3 years). On the other hand, the mean squared error (MSE) improved from the training set (117.5 ± 2.7 years^2^) to the test set (111 years^2^). We also observed that the cardiovascular condition that showed the highest deviation between predicted and biological age, in terms of MAE, was the patients with pacing rhythm (10.5 years), while the patients with prolonged QT-interval had the smallest deviation (7.4 years) in terms of MAE.

This work contributes to existing knowledge of age prediction using deep CNNs on ECGs by showing how a trained model performs on a test set from a separate cohort to that used in the training set.

## 1 Introduction

The electrocardiogram (ECG) was invented by Willem Einthoven in 1901 and since then it has been one of the most important and most frequently used diagnostic tools for cardiovascular diseases. In the 1950s it became possible to convert analog ECG signals to digital ECG signals, this enabled digital interpretation algorithms in the 1960s Smulyan [2019]. These algorithms have generally used rule-based processing techniques to extract features from the ECG in order to classify a large variety of diseases. However, in the last decade, approaches using deep neural networks (DNN) have shown promising performance and present a paradigm shift in how ECGs are being analyzed.

In addition to diagnostic classification, there have been several examples of usage that goes beyond traditional ECG analysis, such as predicting atrial fibrillation in asymptomatic patients Attia et al. [2019b], risk of death Raghunath [2020], gender and age Attia et al. [2019a], Lima et al. [2021], Chang et al. [2022], Ladejobi et al. [2021].

In particular, Attia et al. [2019a] showed that age, predicted by a deep convolutional neural network (CNN), might correlate more with the persons physiological age than the persons biological age. Meaning that, in cases where the predicted age was much higher than the persons biological age, may suggest an underlying disease and might be a biomarker for increased risk of mortality. Furthermore, Lima et al 2021 confirmed, on a separate data set, that the predicted age could be used as a biomarker of the risk of death Lima et al. [2021]. However, previous studies have trained and validated the algorithms on ECG from patients admitted to the same hospitals. This approach might overestimate the performance of the model, and to mitigate this the model should be tested on a separate data set from another hospital. In addition, current studies have either just analyzed predictions from small subsets of patients or looked at high-level risk factors when concluding that CNN-predicted age can be used as a biomarker for disease and mortality. This study therefore set out to train a CNN and validate it on ECGs from a completely separate test set. Furthermore, we will compare the predicted age with the biological age for all ECGs in the test set and categorize the ECGs based on cardiologist-annotated cardiovascular diseases.

## 2 Methods

### 2.1 Data

12-lead ECG recordings from six different open access data bases Reyna et al., Alday et al. [2020], Liu et al. [2018], Tihonenko et al. [2007], Bousseljot et al. [2009], Zheng and et al. [2020] was used to train the proposed model in this study. A seventh data set, collected from another hospital, PTB-XL Wagner et al. [2020] was used as an independent test set. Initially, the training set contained 65900 ECGs and the test set contained 21837 ECGs. After excluding ECGs longer or shorter than 10 seconds and ECGs missing information regarding age or gender the training set contained 59355 ECGs and the test set contained 21748 ECGs. Figure 2 illustrates the exclusion process. The distributions of the patient’s age in the training and test set, after the exclusions, are shown in Figure 1.

**Figure 1:**
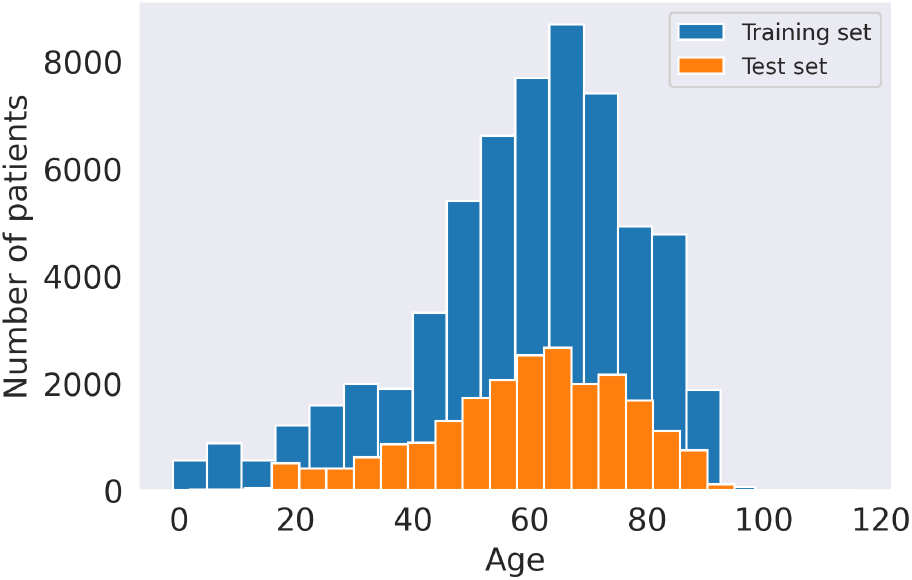
The age distribution of the patients in the training and test set.

**Figure 2:**
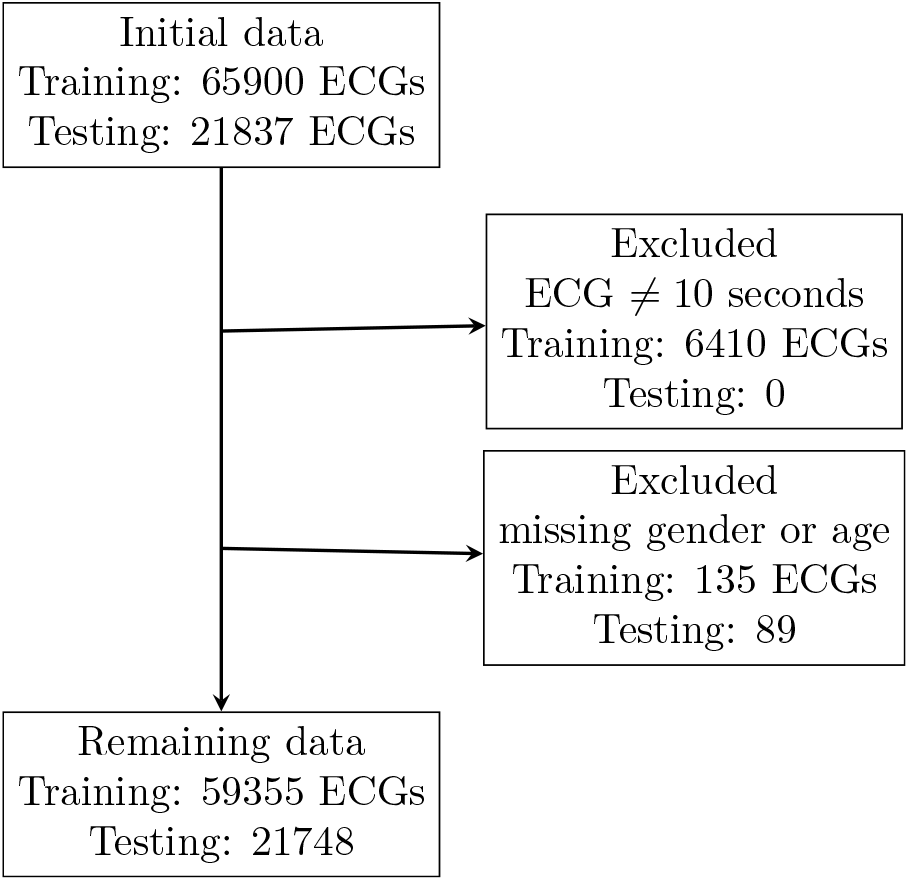
Patients with an ECG recording shorter or longer than 10 seconds or had missing information about gender or age were excluded from the training data.

In addition to the patient’s ECGs, the databases used in this study also contain information about the patient’s age, sex as well as cardiologist-annotated cardiovascular conditions. The age was used as the label to predict by the CNNs, and the cardiovascular conditions were used to categorize the predicted age versus the true age on the test set. Table 1 summarizes the cardiovascular conditions considered in this study and the prevalence of each condition in the test set.

**Table 1:**
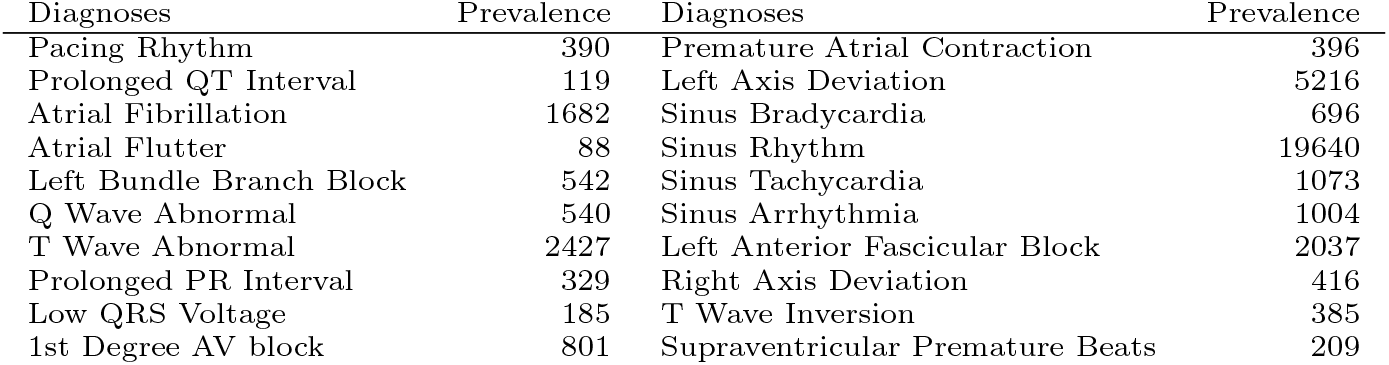
The prevalence of the 20 cardiologist annotated cardiovascular conditions in the test set after exclusions.

### 2.2 Preprocessing

The ECGs were recorded with different electrocardiographs using different sampling frequencies ranging from 256 Hz to 1000 Hz. In this study, we resampled all ECGs to 100 Hz.

### 2.3 Model

Attia et al 2019 proposed a 1-dimensional CNN to predict age from ECGs Attia et al. [2019a]. In this study, we compared the Attia model with a model using an Inception Time architecture Ismail Fawaz et al. [2020]. Both models were trained for 20 epochs with a batch size of 16 using a linear activation function in the last layer and mean squared error as the loss function. The initial learning rate was set to 0.001, but was decreased by a factor of 10 after 10 and 15 epochs.

### 2.4 Validation and testing

The models were first evaluated on the training set using 3-fold stratified cross-validation. The stratification was done based on the patient’s age and gender. Finally, the models were trained on the entire training set and then applied to the test set.

The models were trained using Google Colab with 12 GB GPU and a CPU with 25GB RAM.

## 3 Results

To compare the difference between the Attia model and the Inception Time model we present the difference, in terms of mean absolute error (MAE) and mean squared error (MSE), on the training and test set in Figure 3. The results achieved, using 3-fold cross-validation on the training set, are represented as box and whisker plots, while the performance on the test set is represented with a single point (a star). The Attia model achieved a cross-validated MAE of 8.46 ± 0.09 years and a MSE of 127 ± 2.9 year^2^ on the training set and a MAE of 8.78 years and a MSE of 122.2 year^2^. The Inception model achieved a cross-validated MAE of 7.90 ± 0.04 years and a MSE of 117.5 ± 2.7 year^2^ on the training set and a MAE of 8.3 years and a MSE of 111 year^2^.

**Figure 3:**
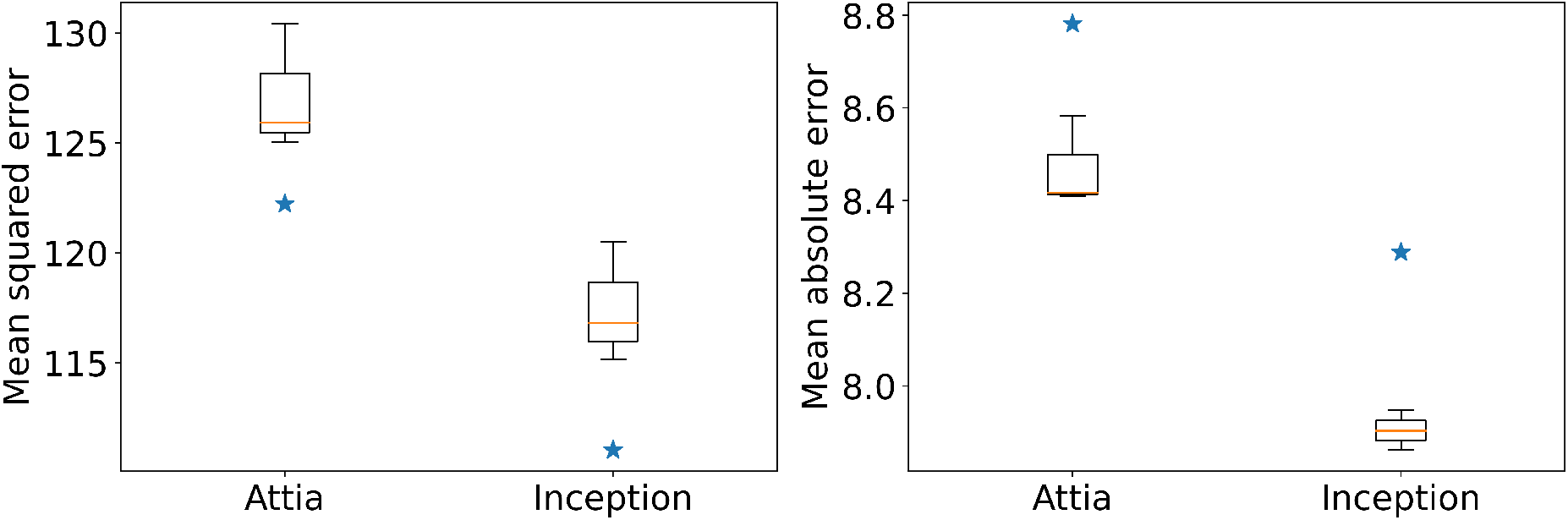
The box and whisker plots represent the mean squared error and the mean absolute error achieved by the Attia model and the Inception Time model using 3-fold cross-validation on the training set. The stars represent the scores obtained on the test set.

Figure 4 provides the relationship between the biological and DNN-predicted age for all of the 21748 patient ECGs in the test set. True versus predicted age by the Attia model are shown in Figure 4a and Inception Time model in Figure 4b. The red line in both figures represents the optimal age prediction, while the green line shows the optimal linear fit between predicted age and true biological age using linear regression.

**Figure 4:**
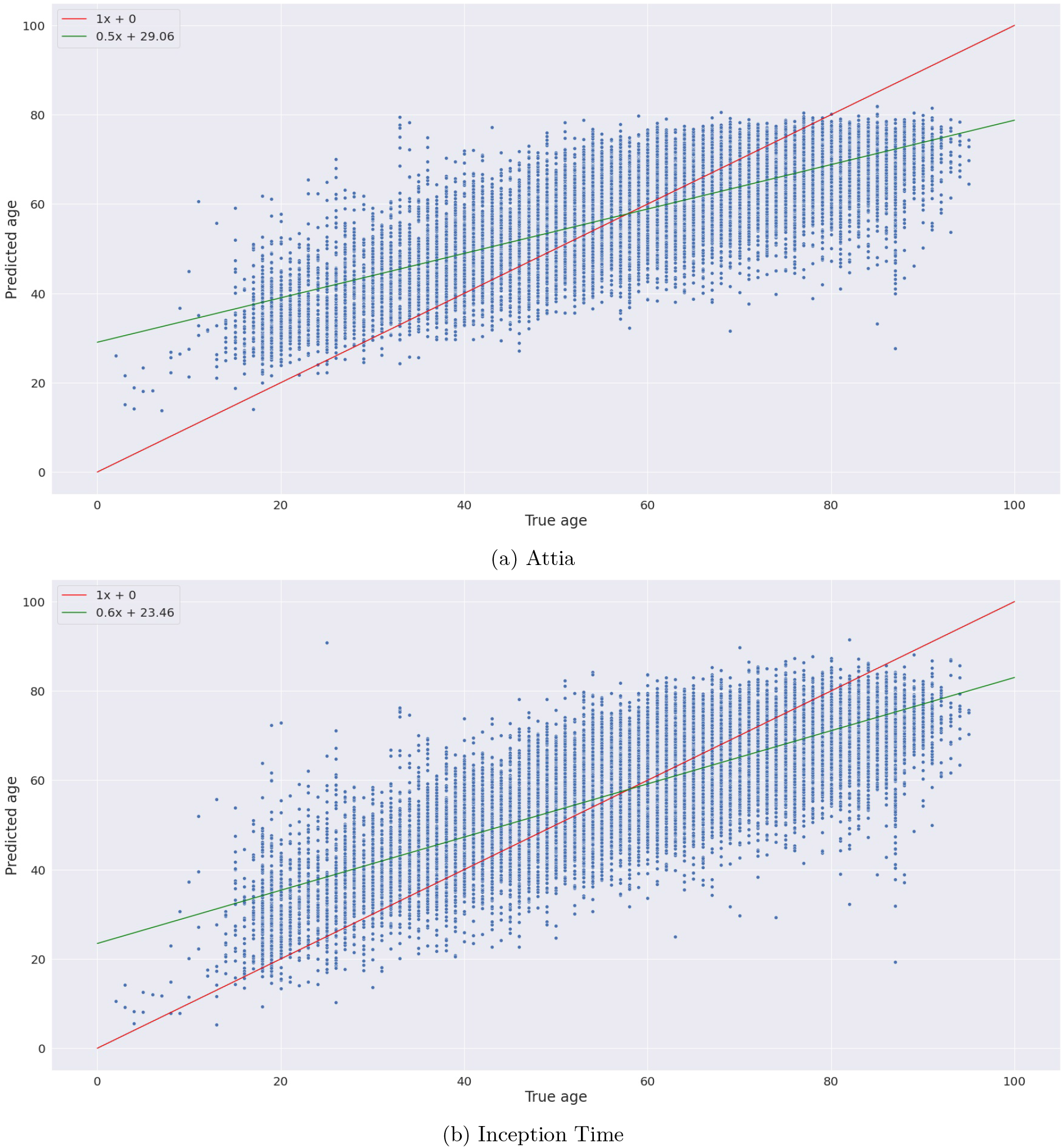
The figures show the relationship between the deep neural network-predicted age and the true (biological) age. The red line shows the best fit for an optimal model, while the green line shows the best linear fit for the current DNN models.

Figure 5 show the CNN predicted age versus the true biological age on the test set, categorized based on the 20 cardiologist-annotated cardiovascular conditions. As in Figure 4 the red lines represent the optimal age prediction, while the green lines show the optimal linear fit between predicted age and true biological age using linear regression. In addition, the MAE for each category is given in the header of each subplot in Figure 5.

**Figure 5:**
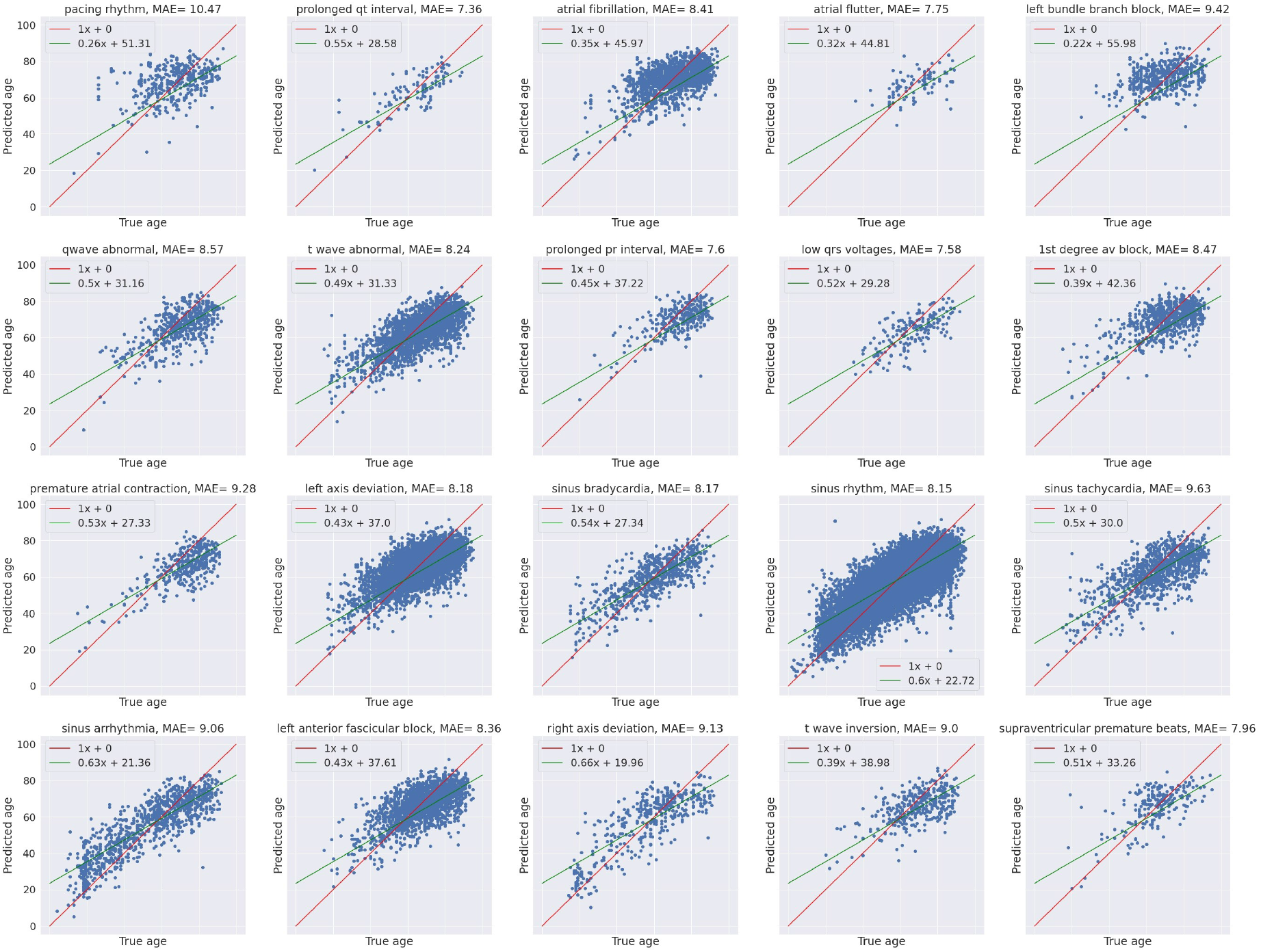
Predicted age from the Inception Time model versus true (biological) age of the patient in the test set. The comparison of predicted and true age are categorized into 20 groups of different cardiovascular conditions.

## 4 Discussion

The findings in this study broadly support the work of other studies in this area, linking ECG with age prediction. In the current study, we trained two CNNs, one proposed by Attia et al 2019 with a second model called Inception Time, to predict age from a persons ECG. Furthermore, we compared the predicted age with the true biological age across all patients in the test set and we also compared them categorized based on cardiologist annotated cardiovascular diagnoses.

Figure 3 shows that the Inception Time model performed significantly better than the Attia model both on the training and the test set. However, from Figure 3 we also see that both models had a significant drop in performance in terms of MAE from training to test set, but it is somewhat surprising that both models also improved the MSE significantly on the test set compared to the training set. The drop in MAE was probably caused by the fact that the models were tested on a data set with patients admitted to a completely different hospital and with a slightly different age distribution, as seen from Figure 1. To understand the improvement in performance in terms of MSE from training to the test set we have to keep in mind that MSE first and foremost punishes prediction outliers. Thus, a possible explanation might be understood by looking at the comparison of true and predicted age in Figure 4 and the age distribution in the train and test set shown in Figure 1. From Figure 4 we see that the biggest discrepancies between true and predicted age are located at each end of the age scale, which is consistent with the reported results in Attia et al. [2019a], and by looking at Figure 1 we see that, in contrast to the training set, there are almost no patients *<* 20 years in the test set, and this might reduce the number of prominent outliers in the test set predictions.

In Figure 5 we have categorized the patients based on cardiologist-annotated cardiovascular conditions, and within each category, we compare the true and predicted age. Contrary to our expectations, the group with the highest MAE (prolonged QT-interval (LQT) MAE= 7.36) was not so different from the group with the lowest MAE (Pacing rhythm MAE= 10.47). In addition, we hypothesized that there would be a linear relationship between the severity of the diagnosis and the degree of misinterpretation in terms of MAE. However, LQT and left bundle branch block (LBBB) are located on each side of the MAE scale, both associated with an increased risk of mortality in contrast to sinus rhythm, for instance, which is considered to be the healthy class in these datasets, but has a MAE less than LBBB and greater than LQT.

A limitation of this study is that the datasets used, both in training and testing, only contain ECGs from patients admitted to the hospital. Even though the ECGs in the training and test set are recorded from different hospitals, from different countries and in some cases with different electrocardiographs, it should still be kept in mind that a large portion of the patients has some sort of disease, since they are admitted to the hospital. In future work, it would be interesting to train a model on a hospital cohort and test it on an independent healthy cohort or vice versa to see how this affects the performance.

## 5 Conclusion

The main goal of the current study was to investigate the performance of a CNN-based age predictor, when tested on a test set from a separate cohort, and compare it to the performance using CV on the training set. The results of this investigation showed that both CNN models tested had a relatively small but significant drop in performance in terms of MAE. This study, therefore, confirms the findings by Attia et al. [2019a], who showed that CNNs could be used to predict a persons age, but also emphasizes the importance of testing such models on separate test sets in order to keep control of possible biases acquired by the model and we therefore strongly suggest that future studies in this field report results on independent test sets in addition to the performance on the training set using some kind of resampling method.

## Data Availability

All data produced are available online at https://physionet.org/ and https://moody-challenge.physionet.org/2021/

https://github.com/Bsingstad/ECG-age

## 6 Code Availability

The code that was used to implement the model and produce the results presented in this paper is hosted on GitHub: not provided yet due to the double-blind review

## Notes

### Competing Interest Statement

The authors have declared no competing interest.

### Funding Statement

This study did not receive any funding

### Author Declarations

The study used ONLY openly available human data that were originally located at https://physionet.org/ and were also used and described in the George Moody Challenge 2021 (https://moody-challenge.physionet.org/2021/)

### Summary of Updates

One of the authors name was spelled wrong. Corrected this

